# Tracking *Candida auris* in Communities via Wastewater: Facility-Level Surveillance and Targeted Sequencing

**DOI:** 10.64898/2026.01.30.26345237

**Authors:** Jingjing Wu, Michael X. Wang, Kaavya Domakonda, Rebecca Schneider, Kirstin Short, Charlene Offiong, Todd J. Treangen, Katherine Ensor, Loren Hopkins, Lauren B. Stadler

## Abstract

*Candida auris* is a multidrug-resistant fungal pathogen that presents substantial challenges for healthcare facilities due to its high mortality rates among vulnerable populations. Six *C. auris* clades have been identified based on their susceptibility to antifungal treatment and environmental stressors. Identifying the circulating *C. auris* clade(s) is critical for understanding transmission and selecting a disease control strategy. To inform targeted implementation of community wastewater monitoring for *C. auris*, samples were collected over 34 weeks from 8 nursing homes and 6 downstream wastewater treatment plants (WWTPs). Detection rates and concentrations of *C. auris* DNA were significantly higher in samples from nursing homes compared to those from WWTPs. Amplicon sequencing methods were developed and applied to characterize the circulating *C. auris* clade in a nursing home wastewater sample. This study demonstrates the utility of wastewater monitoring as a resource-efficient approach for detecting and subtyping *C. auris* in vulnerable communities.

## Introduction

*Candida auris* (*C. auris*) is an emerging fungal pathogen, often resistant to multiple antifungal drugs. First identified in Japan in 2009, it was designated an urgent antibiotic-resistant threat by the Centers for Disease Control and Prevention (CDC) in 2019 ^1,2^. As of December 2023, *C. auris* has been detected in 61 countries. In the United States, clinical cases have increased dramatically – from 51 cases in 2016 to 4,514 in 2023 – with an estimated crude mortality rate of 34% ^3–5^. During the same period, an additional 22,931 screening cases were reported in the U.S., indicating widespread asymptomatic colonization of patients’ skin ^3^.

The transmission of *C. auris* presents significant challenges for healthcare facilities, particularly hospitals and long-term care facilities, due to its persistence in the environment and high mortality rates among vulnerable populations ^6,7^. Colonized patients can contribute to environmental contamination, facilitating further transmission to others within healthcare settings ^3,8^. To date, six distinct *C. auris* clades have been identified, each exhibiting varying levels of multidrug resistance ^6,8–10^. For example, the clade II strains generally exhibit much lower resistance to fluconazole compared to other clades ^6,8,10^, while pan-resistance has primarily been observed in clades I, III, and IV ^10^. Therefore, clade identification is important for informing effective treatment strategies for *C. auris* infections.

Wastewater-based epidemiology (WBE) is an efficient and cost-effective approach for monitoring infectious diseases in populations, including asymptomatic individuals, making it a promising tool for tracking *C. auris* transmission ^11,12^. Several studies have demonstrated the feasibility of detecting *C. auris* in wastewater collected from wastewater treatment plants (WWTPs) and hospitals ^13–17^. However, comparative data between WWTP-level and facility-level surveillance remain limited. For instance, Zulli et al. reported significantly higher detection of *C. auris* in WWTP primary solids when catchment area included healthcare facilities such as hospitals and nursing homes, though their study did not involve direct sampling at the facility level ^14^. Babler et al. observed higher detection rates and concentrations of *C. auris* in hospital grab samples compared to WWTP samples, but the differences were not statistically significant^17^. Furthermore, no study to date has identified the clade of *C. auris* in wastewater samples via direct sequencing.

In this study, we conducted wastewater surveillance for *C. auris* at nursing homes and their corresponding downstream wastewater treatment plants (WWTPs). Detection rates and concentrations were quantified to assess differences in *C. auris* signal between facility-level and community-level sampling locations. To enable clade identification, we developed and validated a tiled amplicon sequencing assay for low-biomass wastewater matrices. In parallel, we developed a bioinformatics pipeline capable of resolving mixed *C. auris* populations and identifying multiple clades within individual wastewater samples.

## Materials and Methods

### Wastewater sample collection, concentration, nucleic acid extraction, and target DNA quantification

To compare *C. auris* detection in facility- and community-level wastewater, samples were obtained once a week from the manhole of 8 nursing homes (NH-A to NH-H) and the influent channel of 6 downstream WWTPs (WWTP-A to WWTP-F, relationships between nursing homes and WWTPs, number of certified beds for each nursing home, and the population served by each WWTP are presented in Table SI.1) across Houston, Texas, from May 11, 2023, to January 4, 2024. For tiled amplicon sequencing assay validation, wastewater samples were collected from WWTPs on March 24 and April 21, 2025, and from NH-D on March 26, 2025.

Samples were collected using refrigerated, time-weighted composite autosamplers and transported to Rice University on ice before concentration and nucleic acid extraction as described by Wu et al. 2025 ^18^. Briefly, each sample was aliquoted into two 50 mL tubes (21008-242, VWR) and centrifuged for 10 minutes at 4,100 g and 4°C to separate the liquid and solid fractions. For the liquid fraction, the supernatant was concentrated by passing through an Electronegative Microbiological Analysis Membrane HA Filter (HAWG047S6, Millipore Sigma). For the wastewater solids, the pellet that remained in the tube was resuspended using 1 mL of lysis buffer from the Chemagic^TM^ Prime Viral DNA/RNA 300 Kit H96 (CMG-1433, PerkinElmer) and transferred into a pre-filled bead-beating tube containing 0.1 mm diameter glass beads. The wastewater solids were used for *C. auris* surveillance via ddPCR, and the liquid fraction was used during the validation of the tiled amplicon sequencing assay to minimize PCR inhibitors. For both fractions, a bead beating step was performed using Mini-Beadbeater 24 (112011, BioSpec) before nucleic acid extraction. Nucleic acid extraction was performed using the ChemagicTM Prime Viral DNA/RNA 300 Kit H96 (Chemagic, CMG-1433, PerkinElmer).

*C. auris* was detected by ddPCR targeting the internal transcribed spacer (ITS2) region. Quantification was performed using ddPCR^TM^ Supermix for Probes (1863024, Bio-Rad) ^19^. The primer and probe sequences, ddPCR reaction assay composition, and thermal cycler conditions are provided in SI 1.3. We followed the EMMI Guidelines ^20^ for quality control and additional information is provided in Table SI.12.

### Assay development for tiled amplicon sequencing

A tiled amplicon sequencing assay was developed to identify the clade(s) of *C. auris* present in a sample (Figure 1a). Nineteen genomes representing all six clades were downloaded from the NCBI database (Table SI.7) ^21^. Core genome alignment was performed using Parsnp (v2.1.2, under default parameters), and a locally collinear block (LCB) was selected based on its length and single-nucleotide polymorphism (SNP) density ^22^. Candidate LCBs for tiled amplicon sequencing were required to have a size between 3,000 and 10,000 base pairs and exhibit high SNP density, defined as the number of SNPs divided by the LCB length. The final candidate LCB, a fragment of chromosome 6 of *C. auris*, has 4,204 base pairs in length and contains 401 SNPs.

**Figure 1.**
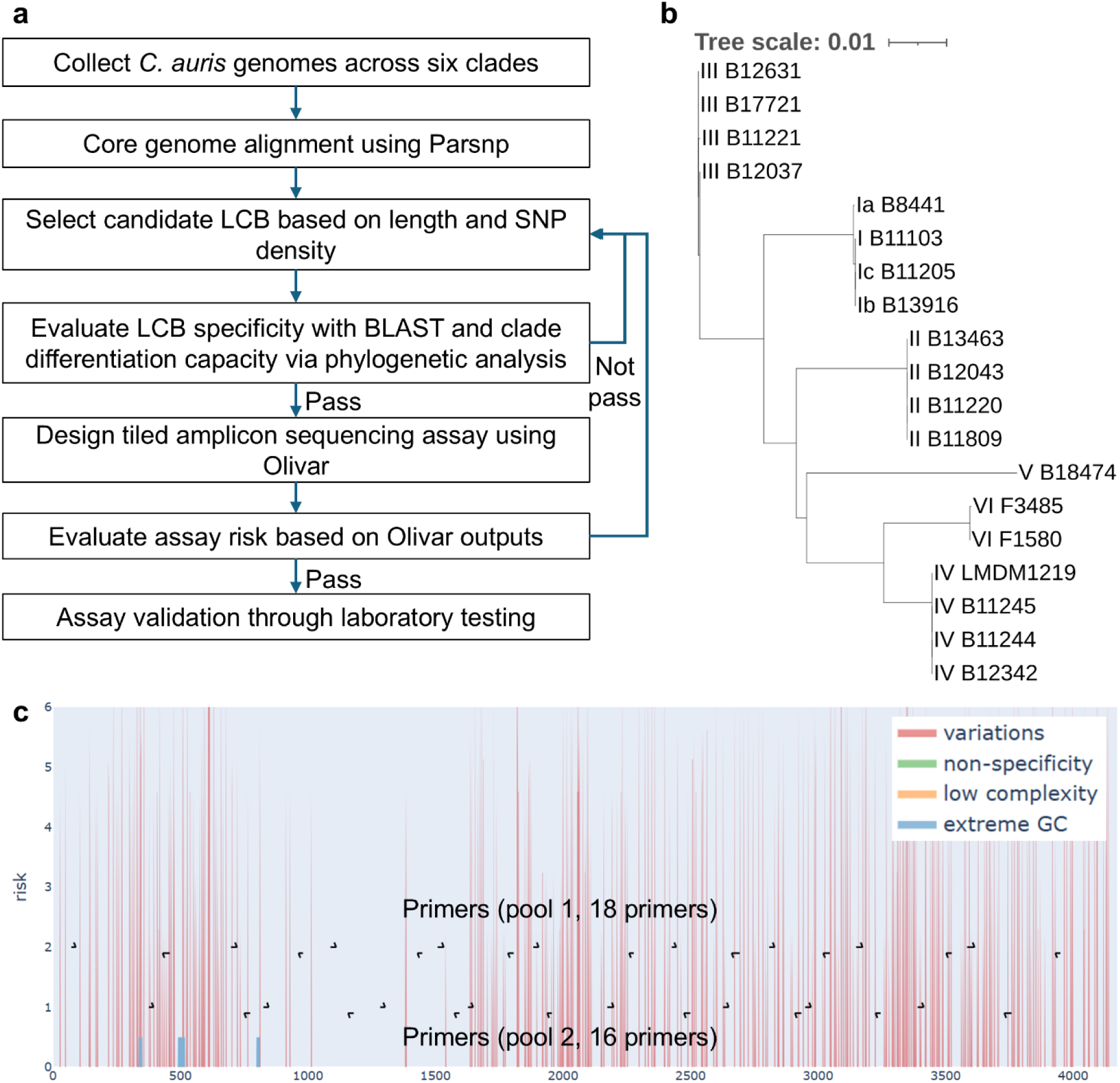
Overview of tiled amplicon sequencing assay design for *C. auris* clade differentiation. **a.** Workflow illustrating the design and validation of tiled amplicon sequencing assay (LCB: locally collinear block; SNP: single nucleotide polymorphism). **b.** Phylogenetic analysis of the selected LCB for *C. auris* tiled amplicon sequencing. Strains from the same clade cluster together, indicating higher intra-clade similarity and inter-clade specificity, which supports the use of the target region for clade differentiation. **c.** Risk landscape of the selected LCB and corresponding primers designed by Olivar. Each risk component is represented in a different color. Primers are assigned to two pools to avoid overlapping amplicons.

To confirm the suitability of the selected LCB for clade differentiation, a phylogenetic tree was constructed using ETE3 (v3.1.3) (Figure 1b) ^22,23^. A consensus sequence was generated using Olivar and BLAST checked against the core_nt database, excluding *C. auris* (taxonomy ID: 498019), to verify the specificity of the target sequence ^24^. Primer design for the tiled amplicon sequencing assay was conducted using Olivar (v1.3.1) ^25^. The desired amplicon size was set to between 252 nt and 420 nt for Olivar, with other input parameters set to default. Assay details can be found in Table SI.8.

### Tiled amplicon sequencing assay validation

We first validated the tiled amplicon sequencing assay using an ATCC standard (MYA-5001, ATCC), which was heat-lysed at 95 °C before use as the template for target sequence amplification. We also validated the assay by spiking the same ATCC standard (MYA-5001, ATCC) into a mixed WWTP influent sample. In addition, we tested the assay on an unspiked nursing home (NH-D) wastewater sample containing endogenous *C. auris*. Lastly, we verified that multiple strains can be identified in an individual sample by spiking pathogen standards of two clades (MYA-5001 for clade II and MYA-5003 for clade IV) into a wastewater sample.

For the spike-in experiment, the ATCC standards were spiked into triplicate 50 mL influent wastewater samples from WWTPs in Houston and gently mixed using a tube rotator (88861051, Fisherbrand) at 10 rpm and 4°C for three hours ^26^. The wastewater samples were concentrated before nucleic acid extraction as described above. The nucleic acid extract was used as the template for both ITS2 quantification using ddPCR (as described above) and tiled amplicon sequencing.

For amplicon sequencing, PCR was performed using Q5^®^ High-Fidelity 2X Master Mix (M0492, New England Biolabs) and primers were added at a final concentration of 200 nanomolar (nM) per primer for each pool. The PCR products for both pools were combined and purified using AMPure XP Beads for DNA Cleanup (A63881, Beckman Coulter Inc.) with a bead-to-sample ratio of 1.8 ^25^. The concentration of purified DNA was quantified using the Qubit dsDNA BR Assay Kit (Q32853, Thermo Fisher) and a Qubit 2.0 fluorometer (Invitrogen), and then diluted to achieve a final concentration of 20 ng/µL in 25 µL before paired-end amplicon sequencing at Azenta (Amplicon-EZ (150-500 bp)).

### Analysis of sequencing data

Paired-end sequencing reads were merged using VSEARCH (v2.15.2) ^27^. Merged reads were quality-filtered using Fastp (v0.23.2) ^28^, with a minimum average read quality score of 30 to reduce sequencing errors and a minimum read length of 236 nucleotides, corresponding to the shortest expected amplicon size. Processed reads were aligned to the reference genome, which was previously generated as a consensus sequence from the selected LCB, using BWA-MEM (v0.7.17) ^29,30^. The resulting SAM files were converted to BAM format, sorted, and indexed using SAMtools (v1.15) ^31^. To enhance alignment accuracy, only reads with an alignment rate greater than 70% to the reference genome were retained. Amplicon primers were trimmed using iVar (v1.4.2) ^32^ before downstream analysis.

Coverage profiles were generated using SAMtools (v1.15) to calculate the breadth of coverage and median sequencing depth. The dominant *C. auris* strain in each sample was identified using Mash (v2.3) ^33^, based on the lowest distance between the sample’s aligned reads (from the BAM file) and the sequences representing each clade. To detect multiple strains within a single sample, haplotype calling was performed on the aligned BAM file using GATK (v4.6.2.0) ^34,35^, with the ploidy set to 2 and soft-clipped bases excluded from analysis. Called variants were compared against clade-specific SNPs, which were derived from an MSA of the representative strains from six clades and the consensus sequence, using the snp-sites (v2.5.1) ^36^. A *C. auris* clade was considered present when its clade-specific SNPs were observed in the variant calls. All Python scripts used in the pipeline are available on GitHub.

### Data analysis

Statistical analysis was conducted using RStudio (2024.12.1, R version 4.4.1). We compared the log-transformed *C. auris* ITS2 concentrations in wastewater (as copies/L-wastewater) measured in samples from nursing homes and downstream WWTPs. The Shapiro-Wilk Test was used to test the normality of the results, and the Mann–Whitney U test was used for the significance test, as the results were not normally distributed.

## Results

### Facility-level and community-level wastewater surveillance for *C. auris*

We compared *C. auris* detection in wastewater samples collected from nursing homes and their corresponding downstream WWTPs (Figure 2). Detection rates were significantly higher in nursing home samples from nursing homes compared to WWTP samples (22.8% vs. 2.5%, p < 0.001). Specifically, *C. auris* DNA was detected in 53 of 232 nursing home samples and in 5 of 196 WWTP samples. Detection across nursing homes was highly variable. For example, NH-D and NH-H, showed high detection rates (80.6% and 48.3%, respectively), whereas others had rates below 30%, and no positive detections were observed in NH-A, NH-C, NH-F, or NH-G. *C. auris* target DNA concentrations also differed significantly between nursing home and WWTP samples (p < 0.001). Nursing home samples had a mean concentration of 10 ^2.83±0.85^ copies/L-wastewater, compared to 10 ^2.44±0.26^ copies/L-wastewater in WWTP samples. This heterogeneity suggests facility-specific differences in *C. auris* transmission or colonization.

**Figure 2.**
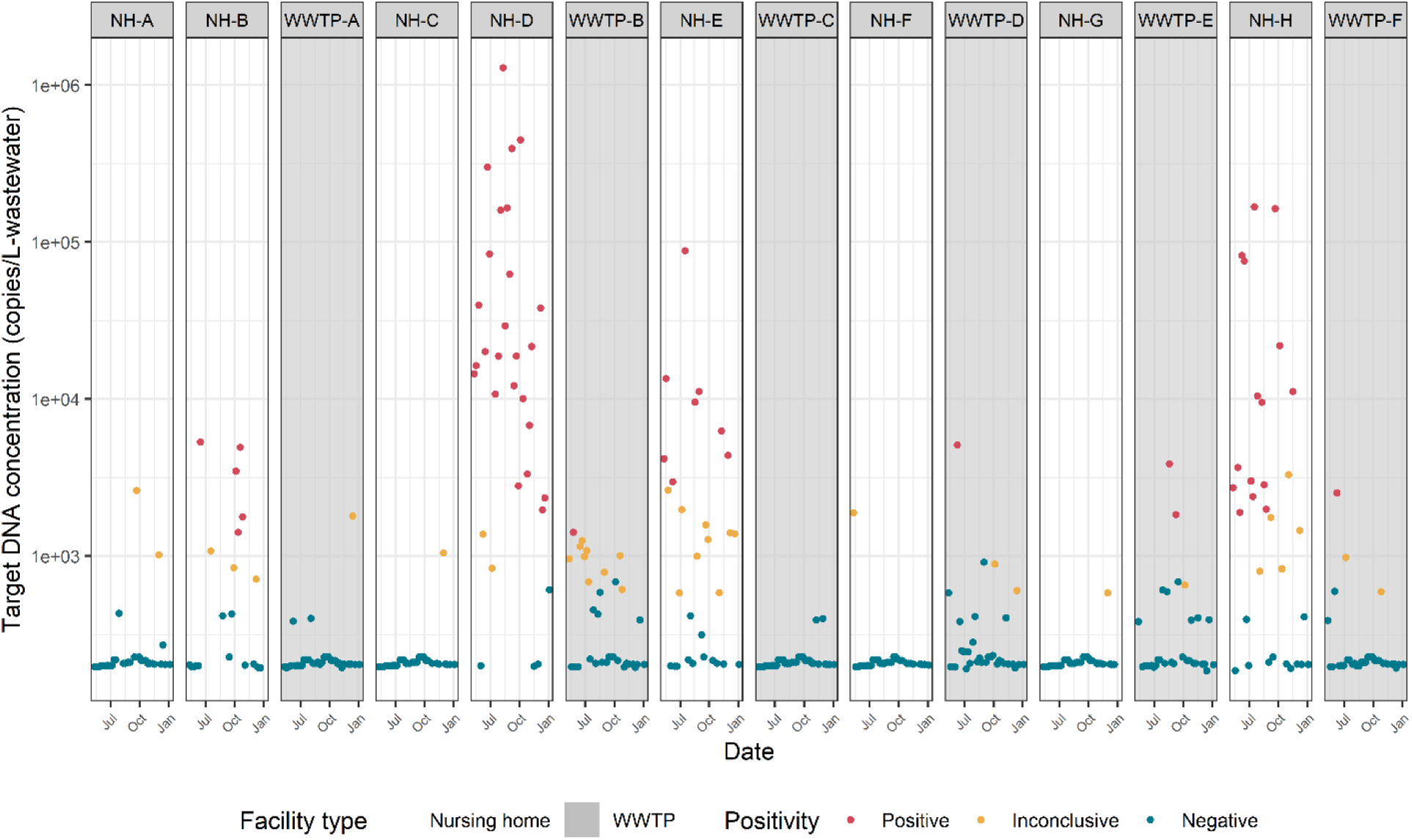
*Candida auris* ITS2 wastewater concentrations in nursing homes and downstream WWTPs. Data from upstream nursing homes (NH) are displayed to the left of their corresponding downstream WWTP. Sample results were classified as follows: positive if target DNA was detected in both duplicates, inconclusive if detected in only one replicate, and negative if not detected in either duplicate.

### Amplicon sequencing and clade differentiation

We validated the amplicon sequencing assay using three sample types: heat-lysed *C. auris* standards, wastewater samples with spiked *C. auris*, and a wastewater sample containing endogenous *C. auris*. Target sequences were successfully amplified in all samples; however, mapping rates, coverage breadth, and sequencing depth varied by sample type (Table 1).

**Table 1.** Results of tiled amplicon sequencing targeting *C. auris*. *C. auris* ITS2 concentrations were measured with ddPCR.

| Type of sample | *C. auris* ITS2 concentration |  | Total number of merged reads | Mapped reads | Mapping rate | Coverage breadth* | Median depth | Spiked standard | Detected clade |
| --- | --- | --- | --- | --- | --- | --- | --- | --- | --- |
|  | Concentration (cp/μL) | Number of positive droplets |  |  |  |  |  |  |  |
| Heat-lysed *C. auris* clade II standard | 139 | 1369 | 391848 | 324813 | 82.89% | 92.50% | 9478 | B11220 (MYA-5001, ATCC) | Clade II |
| Wastewater spiked with *C. auris* clade II standard | 242 | 1780 | 199117 | 106623 | 53.55% | 82.08% | 356 | B11220 (MYA-5001, ATCC) | Clade II |
| Wastewater spiked with *C. auris* clades II and IV standards | 19.8 | 141 | 207996 | 8154 | 3.92% | 88.91% | 103 | B11220 and B11244 (MYA05001 and MYA-5003, ATCC) | Clades II and IV |
| NH-D wastewater | 1.56 | 13 | 279832 | 44 | 0.02% | 31.19% | 0 | - | Clade II |

Compared to heat-lysed standards, wastewater samples generally exhibited lower mapping rates, reduced coverage breadth, and shallower sequencing depths. Among the spiked wastewater samples, a decrease in ITS2 concentration from 242 to 19.8 copies/µL (as measured by ddPCR) resulted in a 92.7% reduction in mapping rate (from 53.55% to 3.92%) and a 71.1% reduction in median sequencing depth (356 to 103). Notably, despite these declines, both samples maintained a coverage breadth exceeding 82%, indicating that genome-wide representation was largely retained.

Clade identification was successful in the heat-lysed standard and both spiked wastewater samples (Table 1; full SNP counts in Table SI.11). Mash analysis identified clade II as the dominant clade in all samples, which was confirmed through clade-specific SNPs analysis. In the sample spiked with both clade II and clade IV standards, SNPs specific to each clade were detected, and was consistent with the clade information provided by the standard manufacturer. These results demonstrate the assay and bioinformatic pipeline’s ability to resolve multiple *C. auris* clades within a single wastewater sample.

The wastewater sample containing endogenous *C. auris* had a low target abundance, with only 1.56 copies/µL and 13 positive droplets detected by ddPCR. Despite the low target concentration, 44 paired-end reads aligned to the reference genome, resulting in a coverage breadth of 31.19%. This limited sequencing output was nevertheless sufficient to enable clade-level identification.

## Discussion

In this study, we demonstrate the feasibility of facility-level wastewater surveillance for *C. auris* using samples collected from nursing homes. Our results show that detection was significantly more sensitive at the nursing home level than at downstream WWTPs, both in terms of detection rates and target DNA concentrations. These findings are consistent with prior studies by Babler et al. (2023) and Zulli et al. (2024), which suggested that wastewater from healthcare facilities contains higher *C. auris* concentrations than WWTP influent ^14,17^.

A key contribution of this work is the direct comparison between nursing homes and WWTP sampling, which revealed statistically significant differences in both detection frequency and target DNA concentrations. These results suggest that healthcare facilities are likely major contributors to *C. auris* levels in wastewater, and that those levels are diluted during transport through the sewer network, reducing detectability at downstream WWTPs. The nursing homes monitored had an average of 161 ± 30 certified beds, while the WWTPs served an average population of 238,396 ± 204,515. This dilution effect has important implications for interpreting WWTP-level surveillance results, as a negative result at the WWTP does not necessarily indicate the absence of the target pathogen in the catchment area, a concern also highlighted previously^37^. For instance, although only one sample from WWTP-B tested positive with a concentration of10^3.15^ copies/L-wastewater, 80.6% of samples from an upstream nursing home (NH-D) were positive, with a maximum concentration of 10^6.11^ copies/L-wastewater – indicating either colonization or ongoing transmission of *C. auris* within the facility. NH-D is a large facility (175 - 200 beds), whereas WWTP-B serves a population of 551,150.

These findings highlight a critical limitation of the current wastewater surveillance strategies, which primarily focus on WWTPs to represent entire sewershed populations ^38^. While this approach may be effective for tracking endemic pathogens, it risks missing low-prevalence threats that disproportionally impact vulnerable populations. Our results underscore the importance of strategic sampling site selection, especially for emerging pathogens such as *C. auris*, where localized outbreaks may not be detectable at aggregate levels.

In this study, we also demonstrate the feasibility of clade-level *C. auris* identification in wastewater using a tiled amplicon sequencing approach. Previous clade differentiation efforts have primarily relied on whole-genome sequencing (WGS) ^9,39,40^, or short tandem repeat (STR) typing ^41^, but these methods have limitations. WGS is resource-intensive and often impractical for wastewater due to low pathogen abundance, DNA degradation and fragmentation, and the complexity of the wastewater matrix ^42^. We also evaluated the STR assay using wastewater with spiked *C. auris* standards and observed poor PCR efficiency and a high rate of length-variant dimers – likely caused by polymerase slippage in the repetitive regions and the presence of inhibitors in wastewater (data not shown) ^43,44^.

Despite lower mapping rates in the raw wastewater sample compared to the spiked samples – an outcome consistent with Wang et al. (2024) – the breadth of sequencing coverage was sufficient to enable clade identification ^25^. This may be attributed to the high density of SNPs in the targeted regions, which enables clade differentiation even with partial amplification. The mapping rate for raw wastewater was only 0.02%, significantly lower than the 4.6%-52.7% range reported Wang et al. (2024) for SARS-CoV-2 tiled amplicon sequencing ^25^. Beyond differences in target pathogen, assay design, and wastewater variability, the primary contributor to this discrepancy is likely the initial target concentration: while Wang et al. reported ddPCR-based viral concentrations ranged from 28.1 to 106.9 cp/µL, our endogenous *C. auris* ITS2 concentration was only 1.56 cp/µL. Furthermore, because the ITS2 region is located within the tandemly repeated rDNA cluster, its apparent concentration may overestimate the true *C. auris* concentration ^45^. Despite these challenges, our results confirm that clade identification is feasible even in low-abundance samples.

The limited sequencing coverage depth and breadth observed in the raw wastewater sample highlight opportunities to improve the tiled amplicon sequencing pipeline. Increased sequencing depth will be particularly important for reliably identifying multiple clades within a single sample. Additionally, improvements to the clade identification pipeline could expand beyond reliance on clade-specific SNPs alone. Integrating the co-occurrence of mutations within individual reads and incorporating read-level mutation depth could enable more robust clade identification and even clade abundance estimation. Other improvements include developing ddPCR assays targeting single copy genomic regions to improve quantification accuracy, and establishing the detection limits of this clade-level identification workflow.

The tiled amplicon sequencing assay developed and validated here represents a valuable tool for public health surveillance by enabling clade-level identification of *C. auris* using wastewater. For example, our sequencing analysis of the nursing home wastewater sample revealed the presence of Clade II *C. auris*, which is typically associated with ear infections and colonization and tends to be more susceptible to antifungal drugs ^10,46,47^. In contrast, detection of Clades I, III, or IV, which are associated with a higher risk of pan-resistance and invasive infection, would raise more serious public health concerns ^10^. The ability to rapidly identify and differentiate these clades from wastewater provides critical information to guide targeted response efforts.

Future research should focus on pairing wastewater surveillance with point prevalence at facilities. This would enable a comparison of wastewater concentrations with information on active infections, colonization, and potential environmental sources. Shedding estimates for colonized and infected individuals could also inform the interpretation of wastewater surveillance results, and wastewater could be used to estimate shedding if performed in parallel with point prevalence surveys. A potential limitation of *C. auris* wastewater monitoring in healthcare facilities is the reduced sensitivity for patients who rely on adult diapers. Wastewater monitoring could be applied to assess strategies for preventing transmission of *C. auris* from colonized and infected individuals, and for early detection of transmission events in facilities housing vulnerable populations.

## Data Availability

All data produced in the present study are available upon reasonable request to the authors

## Acknowledgments

This research was supported by funds from the Centers for Disease Control and Prevention (NI50CK000557). We thank Houston Water for their assistance in sample collection. We thank Lauren Bauhs and Robert Campos for their help with wastewater processing.

## Notes

### Competing Interest Statement

The authors have declared no competing interest.

