## Supplementary material for "Tracking *Candida auris* in Communities via Wastewater: Facility-Level Surveillance and Targeted Sequencing": SI

### SI 1. Materials and Methods

#### SI 1.1 Wastewater sample collection

**Table SI.1:** Relationship between nursing home and downstream wastewater treatment plant (WWTP)

| Nursing home | Size * | Corresponding downstream WWTP | Population served |
|---|---|---|---|
| NH-A | Medium | WWTP-A | 109,414 |
| NH-B | Large | WWTP-A | 109,414 |
| NH-C | Medium | WWTP-B | 551,150 |
| NH-D | Large | WWTP-B | 551,150 |
| NH-E | Small | WWTP-C | 70,900 |
| NH-F | Large | WWTP-D | 124,000 |
| NH-G | Medium | WWTP-E | 293,227 |
| NH-H | Small | WWTP-F | 97,918 |

* The size of a nursing home is determined based on the number of beds. Small: <=125 beds, medium: 126-175 beds, and large: >175 beds.

#### SI 1.2 Wastewater sample processing

For spiked wastewater samples, the *C. auris* standards were diluted and mixed before being spiked into 50 mL influent wastewater samples from WWTPs in Houston. The spiked wastewater samples were gently homogenized using a tube rotator (88861051, Fisherbrand) at 10 rpm and 4ºC for 3 hours [1]. The concentration methods were previously described and summarized as follows [2–4]. The wastewater sample was aliquoted into 50 mL centrifuge tubes. Tubes containing wastewater were centrifuged at 4,100 g and 4ºC for 10 minutes to separate liquid and solid fractions. For the liquid fraction, the supernatant was carefully poured into an

MF 3, 300ml Magnetic Filter Holder with lid kit (20030001, Sterlitech), which was connected to a Multi-Vac 600-MS Manifold (180600-01, Sterlitech) and a Rocker 800 Oil Free Laboratory Vacuum Pump (167800, Sterlitech) system. The Electronegative Microbiological Analysis Membrane HA Filter (HAWG047S6, Millipore Sigma) was placed into the manifold system before sample addition. To enhance adsorption, 1 mL of 1.25 M $MgCl_2 \cdot H_2O$ (M0250, Sigma Aldrich) solution was added to the sample, gently mixed using the pipette tip, and allowed to sit for 5 minutes before vacuum filtration. After filtration, the membrane filter was folded and transferred into a bead-beating tube pre-filled with 0.1 mm diameter glass beads and 1 mL of lysis buffer from the chemagic™ Prime Viral DNA/RNA 300 Kit H96 (CMG-1433, PerkinElmer).

For the wastewater solids, following supernatant removal, the remaining pellet was resuspended in 1 mL of lysis buffer from the same Chemagic™ kit and transferred into a bead-beating tube pre-filled with 0.1 mm diameter glass beads. All bead-beating tubes were processed using the Mini-Beadbeater 24 (112011, BioSpec) for 1 minute at 3,500 oscillations/m, repeated twice with a 2-minute interval on ice between runs. Following bead beating, tubes were centrifuged at 17,000 g for 5 minutes at 4°C, and 300 µL of lysate was used as input to nucleic acid extraction using Chemagic™ Prime Viral DNA/RNA 300 Kit H96 (Chemagic, CMG-1433, PerkinElmer), following the manufacturer's protocol. Extracted nucleic acids were eluted in 50 µL of sterile, nuclease-free water and stored at 4°C for no more than 48 hours before being used for PCR quantification or sequencing.

The concentrations of *C. auris* ITS2 region were quantified using ddPCR™ Supermix for Probes (1863024, Bio-Rad). Droplet generation was performed using an Automated Droplet Generator (1864101, Bio-Rad) and RT-ddPCR was completed using a C1000 ThermalCycler

(1851197, Bio-Rad) and QX600 AutoDG Droplet Digital PCR System (12013328, Bio-Rad). Results were analyzed using QuantaSoft v1.7.4 software.

For amplicon sequencing, the PCR was performed using Q5® High-Fidelity 2X Master Mix (M0492, New England Biolabs) and designed primers at a final concentration of 200 nanomolar (nM) per primer. The amplification was performed using each pool of primers separately, and the PCR products for both pools were combined before purification using AMPure XP Beads for DNA Cleanup (A63881, Beckman Coulter Inc.), following the manufacturer's protocol [5]. 80 µL of PCR products were mixed with 144 µL of AMPure XP Beads for DNA Cleanup (A63881, Beckman Coulter Inc.), washed twice with 70% ethanol, and eluted into 40 µL of nuclease-free water (AM9930, Thermo Fisher). The purified DNA products were normalized to 20 ng/µL in 25 µL using Qubit dsDNA BR Assay Kit (Q32853, Thermo Fisher) and a Qubit 2.0 fluorometer (Invitrogen) before being submitted for paired-end amplicon sequencing at Azenta (Amplicon-EZ (150-500 bp)).

**Table SI.2:** Wastewater sample concentration factor

| | **Volume** | | **Concentration factor** |
|---|---|---|---|
| Sample volume: | 50 | mL | |
| Lysis buffer added | 1000 | µL | 50 |
| Added to Chemagic | 300 | µL | |
| Elution volume: | 50 | ul | 6 |
| | | **Total:** | 300 |

**SI 1.3: ITS2 ddPCR assay and thermal cycling conditions**

**Table SI.3:** ddPCR assays and positive standards used for *C. auris* ITS2 region quantification [6]

| Assay name | Sequence (5'-3') |
|---|---|
| Forward Primer | CAGACGTGAATCATCGAATCT |
| Reverse Primer | TTTCGTGCAAGCTGTAATTT |
| Probe | SUN/AATCTTCGC/ZEN/GGTGGCGTTGCATTCA/3IABkFQ |
| Amplicon length | 135 |
| Gblock sequence * | TTTCGTGCAAGCTGTAATTTTGTGAATGCAACGCCACCGCGAA<br>GATTGGTGAGAAGACATCACGCTCAAACAGGCATGCCTTGGG<br>GAATACCCCAAGGCGCAATGTGCGTTCAAAGATTCGATGATTC<br>ACGTCTGCAAGTTTGCACCAGTGACTATGAATAACGATGAAGC<br>TGTGGATTTTGCTAGTGAGGTTGCAAAAGAATTATTTGGCGAA<br>AAAAATTGTGAATTTAATCATCGTCCTTTAATGGCAAGTGAGG<br>ATTTTGGATTAGCGTACTGGAAAGGGAAAGTCAGCTTTACGGT<br>TCCTTTGACGGTGCGATGAAGTTTATCAAAGGTGACGCCATTG<br>CCGGTATCATTATCATCTTTGTGAACTTTATTGGCGGTATTAAGA<br>AATTTGTCATGCTCTTCAGGAGATGAAACGAAATGTCGTAAAA<br>TTGGACAATCTTCAGCAACAAATTGTCCAAAGACTCGCAAAG<br>AAAGTTTTGGTAGAACATGTGA |

* The gblock sequence includes target sequences of other pathogens that are multiplexed with *C. auris* in our lab

**Table SI.4:** Final concentrations of the primer-probe mix

| Assay component | Final concentration (µM) | 20x concentration (µM) |
|---|---|---|
| Forward primer | 0.9 | 18 |
| Reverse primer | 0.9 | 18 |
| Probe | 0.25 | 5 |

**Table SI.5:** Reaction composition for the ddPCR assay

| Reagent | Volume (µL) |
|---|---|
| ddPCR Supermix (No dUTP) | 10 |
| Primer-Probe mix of each target (40x) | 1 |
| RNase/DNase-free water | 1 |
| DNA template | 10 |

*Final concentrations in reaction: 0.25µM (probe) and 0.9µM (forward and reverse primers)

**Table SI.6:** Thermal cycling conditions for the ddPCR assay

| Cycling step | Temperature °C | Time | Number of cycles |
|---|---|---|---|
| Enzyme activation | 95 | 10 min | 1 |
| Denaturation | 94 | 30 sec | 40 |
| Annealing/Extension | 60.1 | 60 sec | |
| Enzyme deactivation | 98 | 10 min | |
| Hold (optional) | 4 | Infinite | 1 |

#### SI 1.4 Assay development for tiled amplicon sequencing

**Table SI.7:** List of *Candida auris* (*C. auris*) genomes used for tiled amplicon sequencing assay design and clade identification.

| Strain name | Clade | Accession number |
|---|---|---|
| B11103 | I | GCA_031359945.2 |
| B8441 | Ia | GCA_002759435.3 |
| B13916 | Ib | GCA_016772235.1 |
| B11205* | Ic | GCA_016772135.1 |
| B12043 | II | GCA_016495645.1 |
| B13463 | II | GCA_016495665.1 |
| B11809 | II | GCA_016495685.1 |
| B11220* | II | GCF_003013715.1 |
| B17721 | III | GCA_016772175.1 |
| B12631 | III | GCA_016772195.1 |
| B12037 | III | GCA_016772215.1 |
| B11221* | III | GCA_031357565.2 |
| B11245 | IV | GCA_008275145.1 |
| B12342 | IV | GCA_016772155.1 |

| | | |
|---|---|---|
| B11244* | IV | GCA_031357835.2 |
| LMDM 1219 | IV | GCA_041381755.1 |
| B18474* | V | GCA_016809505.1 |
| F1580 | VI | GCA_032714025.1 |
| F3485* | VI | GCA_032715285.1 |

* Sequences used to represent each strain for clade identification. One representative strain per clade was selected based on the sequence conservation within clades in the target region (see Figure 1b)

#### SI 1.5 Tiled amplicon sequencing assay and thermal cycling conditions

**Table SI.8:** *C. auris* tiled amplicon sequencing assay

| Amplicon ID | Pool | Forward primer | Reverse | Start* | End* |
|---|---|---|---|---|---|
| Cauris_1 | 1 | GCCCGAGTTTCCCGTGT | CAATTCCCGTTTTGTCATAAAATTGACT | 74 | 459 |
| Cauris_2 | 2 | GGCAGTGGGGGTGGTGAA | AGCCTAGAGTGTCTAGAGTGTGT | 378 | 774 |
| Cauris_3 | 1 | ACTACCCAACATCATCACCCTAC | TGCCCACGGCAACTCCT | 700 | 981 |
| Cauris_4 | 2 | CAGGGTCCAAGTAGTAGGCAG | GACGTTCTTTTGACGTTCCTCC | 827 | 1180 |
| Cauris_5 | 1 | GCTGGGTCAATGTCACGGTATC | GGATGTGCCCCTCTCCGAA | 1090 | 1449 |
| Cauris_6 | 2 | ACAGGCACGTAGAGCTGGT | CTGGACCGCACGGGTATAAA | 1285 | 1595 |
| Cauris_7 | 1 | AGCTTAGGCATTGTACTTTGTGTG | TTGTTTGGTGACGTTATGCGTT | 1510 | 1807 |
| Cauris_8 | 2 | GGTGACTTTTGAAGCTGGCG | CTTCCAGGGCCCACAGTC | 1629 | 1957 |
| Cauris_9 | 1 | AAGGGCTCTCCTTCTCTCAGT | TTGCCAGCTTGCCTCGAT | 1885 | 2278 |
| Cauris_10 | 2 | CAGTTTCCAGTATGAACATTGCCAT | GAATGCAACGGAAAAATCACTGAAA | 2175 | 2500 |
| Cauris_11 | 1 | GCCCAAGTCGGGGCCTT | ACAATGTCTACTTTAAGAGGAAGTGTTTTAA | 2428 | 2693 |
| Cauris_12 | 2 | ACGTCTCGTTGTATCGACATGA | GGTTTGTGTATTTGGTGTTGCTGTA | 2629 | 2935 |
| Cauris_13 | 1 | ACATTGTTCCAAGATGCCCCT | CACCCTCCTCAATTCTTCCAATATTT | 2810 | 3047 |

| | | | | | |
|---|---|---|---|---|---|
| Cauris_14 | 2 | GGTCTGGACTTGAGATCGTTGTA | GAACTCGTCAACCCGGTGAA | 2951 | 3246 |
| Cauris_15 | 1 | ACTTTCCACTTTTCTGGAGAGTGAA | TGGCAAGGTTCAAGAGACAACT | 3151 | 3525 |
| Cauris_16 | 2 | GGTTTGCCGATTGAACATACTTCT | ACCTTATAATTTGAGTTCCCAAAGTGAA | 3392 | 3759 |
| Cauris_17 | 1 | CTTTTCGTTCTCTTCTCACTAAAACTACT | CTCCAGTGGATCACCTGCTT | 3586 | 3951 |

*Start and end represent the coordinates of the first and last base pairs of the amplicon on the reference genome.

**Table SI.9:** Reaction composition for PCR (50 µL reaction)

| Reagent | Volume (µL) | Final concentration |
|---|---|---|
| Q5® High-Fidelity 2X Master Mix | 25 | 1X |
| Premixed primers (1 µM for each primer) | 10 | 200 nM |
| RNase/DNase-free water | 7 | - |
| DNA template | 8 | - |

*Final concentrations in reaction: 0.25µM (probe) and 0.9µM (forward and reverse primers)

**Table SI.10:** Thermal cycling conditions for the PCR assay

| Cycling step | Temperature °C | Time | Number of cycles |
|---|---|---|---|
| Enzyme activation | 95 | 10 min | 1 |
| Denaturation | 94 | 30 sec | 40 |
| Annealing/Extension* | 52.4 / 51.9 / 51.2 / 50.6 | 60 sec | |
| Enzyme deactivation | 98 | 10 min | |
| Hold (optional) | 4 | Infinite | 1 |

*50 µL reaction was divided into four aliquots of 12 µL each. Reactions were run at four different annealing temperatures to optimize the amplification efficiency across primer sets.

#### SI 1.6 Quality control measures and the theoretical limit of detection (LOD) calculation

The quality control measures and the calculation of theoretical LOD are described by Low et al. (2022) [3] and Wolken et al. (2023) [7] and are summarized briefly as follows. Two bead tubes containing glass beads and lysis buffer were included as extraction negative controls. In addition, each ddPCR quantification plate included at least two no-template controls (NTCs) with RNase/DNase-free water and two positive controls using gBlock Gene Fragments (IDT, USA; sequence provided in SI 1.3).

An acceptable total droplet count of at least 10,000 was established for all sample wells as recommended by the manufacturer. Each method's theoretical limit of detection (LOD) for ddPCR was determined as three positive droplets per well plus the maximum number of positive droplets among the negative controls. The theoretical LOD was converted to copies per µL of DNA template and copies per liter of wastewater based on the estimated droplet volume (0.86 nL), the number of total droplets, the volume fraction of DNA template within a droplet (10/22), and the concentration factor during sample processing. The equations used for the LOD calculation are presented in SI. Eq 1-3. For samples with no positive droplet detections, the concentration was imputed as half the concentration corresponding to one positive droplet.

<u>Theoretical Limit of Detection (LOD) Calculation Equations</u>

**SI. Eq 1:**

$$LOD_{droplet} = 3 + maximum\ number\ of\ positive\ droplets\ across\ all\ process\ blanks^{*}$$

* Process blanks include: two extraction blanks, and no less than two no template controls per plate included in ddPCR quantification.

**SI. Eq 2:**

$$LOD_{\mu L-DNA\ template} = \frac{LOD_{droplet}}{0.86\frac{nl}{droplet} \times n\ total\ droplets\ in\ each\ well\ \times \frac{10}{22} fraction\ of\ template\ within\ droplet}$$

**SI. Eq 3:**

$$LOD_{L-wastewater} = LOD_{\mu L-DNA\ template} \times \frac{1}{Concentration\ factor\ (300)} \times \frac{1000000\ \mu L}{1\ L}$$

#### SI 2. Results

**Table SI.11:** Number of clade-specific SNPs identified in heat-lysed *C. auris* and wastewater samples, along with the detected clade for each sample.

| **Type of sample** | **Spiked standard** | **Number of specific SNPs that support each clade** | | | | | | **Detected clade** |
|---|---|---|---|---|---|---|---|---|
| | | I | II | III | IV | V | VI | |
| Heat-lysed *C. auris* standard | B11220 (MYA-5001, ATCC) | 0 | 26 | 0 | 0 | 0 | 0 | Clade II |
| Wastewater spiked with one clade of *C. auris* standard | B11220 (MYA-5001, ATCC) | 0 | 25 | 0 | 0 | 0 | 0 | Clade II |
| Wastewater spiked with two clades of *C. auris* standard | B11220 and B11244 (MYA05001 and MYA-5003, ATCC) | 0 | 25 | 0 | 16 | 0 | 0 | Clades II and IV |
| Raw wastewater from NH-D | - | 0 | 5 | 0 | 0 | 0 | 0 | Clade II |
